# A simple arithmetic rationale for crushing the epidemic curve of Severe Acute Respiratory Syndrome Coronavirus 2 (SARS-CoV-2) instead of flattening it

**DOI:** 10.1101/2020.05.06.20093112

**Authors:** Gerry F. Killeen

## Abstract

Countries with ambitious strategies to *crush the curve* of their epidemic trajectories, to promptly eliminate SARS-CoV-2 transmission at national level, include China, Korea, Japan, Taiwan, New Zealand and Australia. In stark contrast, many of the European countries hit hardest over the last two months, including Italy, Spain, France, Ireland and the United Kingdom, currently appear content to merely *flatten the curve* of their epidemic trajectories so that transmission persists at rates their critical care services can cope with. Here is presented a simple set of arithmetic modelling analyses that explain why preferable crush the *curve strategies*, to eliminate transmission within months, would require only a modest amount of additional containment effort when compared to *flatten the curve* strategies that allow epidemics to persist at a steady, supposedly manageable level for years, decades or even indefinitely.

---

Most cases of the severe acute respiratory syndrome coronavirus 2 (SARS-CoV-2) are relatively mild or even asymptomatic,^1,2^ and transmission can occur through such subtle mechanisms as droplets generated while speaking^3^ and persistence on contaminated surfaces.^4^ Reactive containment interventions against SARS-CoV-2, based on testing and contact tracing, are therefore unlikely to succeed as a stand-alone containment measures.^1,2^ Furthermore, it remains to be seen whether any sufficiently effective new vaccines or drugs can be developed, evaluated and made available globally in sufficient quantities soon enough to avert the worst consequences of the ongoing SARS-CoV-2 pandemic.^5^ In the meantime, the only effective intervention options available to governments are various presumptive social distancing, hygiene and quarantine measures, enforced variations of which are often referred to as *lock down*.

However, different countries appear to be applying these behavioural interventions to achieve quite distinct targets for their epidemic trajectories.^6^ Examples of countries with ambitious strategies to *crush the curve^7^* of their epidemic trajectories, to promptly eliminate SARS-CoV-2 transmission at national level, include China, Korea, Japan, Taiwan, New Zealand and Australia. In stark contrast, many of the European countries hit hardest over the last two months, including Italy, Spain, France, Ireland and the United Kingdom, currently appear content to merely *flatten the curve^8^* of their epidemic trajectories so that transmission persists at rates their critical care services can cope with. Here is presented a simple arithmetic rationale for why preferable *crush the curve* strategies, to eliminate transmission within months, would require only a modest amount of additional containment effort when compared to *flatten the curve* strategies that allow epidemics to persist at a steady, supposedly manageable level for years, decades or even indefinitely.^2,9^

Much can be learned by simply examining the targets for the two alternative strategies, relative to the starting point, expressed in terms of the reproductive number of the virus (*R*) or number of new infections arising from any initial infection over its full duration. An epidemic curve which has been exactly flattened, so that the rate of incidence of new infections remains constant (*R*_0_=1.0), represents a tipping point in the struggle to contain SARS-CoV-2. Once the reproductive number has been pushed below this critical threshold, even modest further reductions achieve a snowball effect that crushes the epidemic curve by progressively accelerating progress towards elimination of local transmission (Figure 1A).

**Figure 1.**
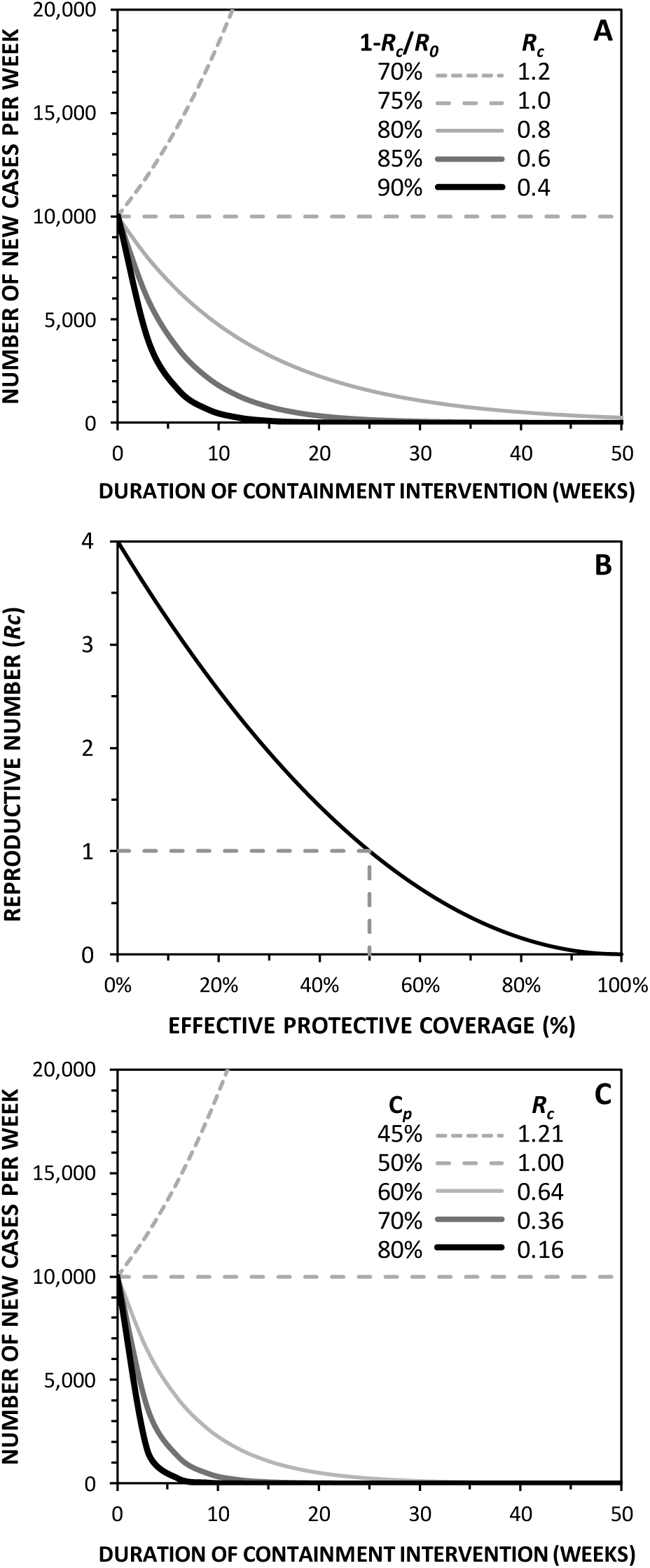
The effects of varying levels of containment effectiveness upon the expected subsequent trajectories of a SARS-CoV-2 epidemic. It was assumed that the epidemic had reached an incidence rate of 10,000 new infections per week at the point when a suite of presumptive, population-wide preventative behavioural interventions (often referred to as *lock down* if enforced) were introduced, with an initial preintervention reproductive number of 4 new infections per existing infection (*R*_0_=4.0). **A**: Controlled reproduction numbers (*R_c_*) and incidence rate trajectories expressed as functions of overall reduction of transmission rate (1-*R_c_*/*R*_0_). **B**: Controlled reproductive numbers (*R_c_*) expressed as a function of mean effective protective coverage of individuals with interventions to prevent exposure behaviours (*C_p_*). **C**: Controlled reproductive numbers (*R_c_*) and incidence rate trajectories expressed as functions of effective protective coverage of individuals with interventions to prevent exposure behaviours (*C_p_*).

For example, if we assume an approximate median between the most conservative (lowest) and insightful (highest) estimates for the reproductive number of the virus under the unconstrained conditions before interventions were introduced (*R*_0_) at the outset of well-documented outbreaks, a baseline value of 4.0 seems as reasonable as any.^2^ From this assumed starting point, a country that contains its epidemic sufficiently to flatten the curve to a plateau, so that the rate of incidence of new infections remains constant, would have achieved a controlled reproductive number (*R_c_*) of exactly 1.0 (Figure 1A). Relative to where that country started, this minimum containment level required to prevent the epidemic from growing further would represent a 75% reduction of transmission. Countries like Ireland, France, Spain, Italy and the United Kingdom, where daily incidence rates are now slowly falling (*R_c_*<1.0), so their epidemics could slowly fizzle away if current measures were maintained, may well have achieved 80% suppression of transmission. If sustained, current measures in these countries could be expected to see their incidence rates shrink by 98% but not disappear over the course of a year (Figure 1A). While this is a considerable achievement, it begs the question why these countries would not build upon their successes by further improving their epidemic response intervention packages to steepen the downslope they now find themselves on and then pursue the sequential goals of elimination and exclusion? By improving the impact of control efforts beyond the initial flattening threshold of 75% transmission reduction, so that a further 10% of transmission is prevented, would result in an overall transmission reduction of 85% and an epidemic that would contract by 40% (*R_c_*=0.6) every 3 weeks (Approximate mean duration of infection^2^) before petering out after a year (Figure 1A). Squeeze transmission down by just another 5% overall (90% reduction, *R_c_*=0.4) and local transmission may collapse within 30 weeks (Figure 1A).

While these levels of transmission suppression may sound very high, several countries (notably China,^10^ which was hit first and without warning at the outset) have achieved controlled reproductive numbers and incidence shrinkage rates in this approximate range, so they are now approaching their elimination targets. Furthermore, such impressive reductions of transmission rate and relatively rapid escape trajectories from self-sustaining local transmission may be far easier to rationalize in simple arithmetic terms by considering two important, intuitive and encouraging non-linearities of pathogen outbreak and containment dynamics: (1) Transmission requires exposure behaviours by two individuals, so transmission varies in proportion to the square of the relative rates of those preventable exposure behaviours, and (2) Even modest acceleration of proportional decay rates can dramatically curtail the length of time it takes for them to approach zero.

Transmission from one individual to another requires exposure behaviours by two people who interact through direct contact or through shared spaces, surfaces and objects. Transmission rate is therefore proportional to the product of their individual exposure behaviour rates, which in turn depends on limitations of intervention coverage and effectiveness once containment measures are introduced. Correspondingly, the reproductive rate achieved by such control measures may be calculated as a simple squared function of the gap in the population mean effective protective coverage (*C_p_*) for a preventative intervention suite:

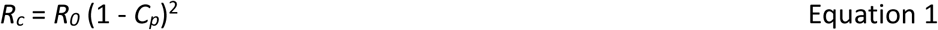

where effective protective coverage is the product of the population mean compliance coverage (*C*) and effective personal protection (*ρ*) achieved by compliant individuals (*C_p_* = *ρ C*).^2^ This intuitively non-linear relationship indicates that proportional impacts upon transmission may be reasonably expected to exceed effective protective coverage (Figure 1B), so thresholds for flattening or crushing the epidemic curve may be achieved more readily than would otherwise be envisaged (Figure 1C). When intervention effectiveness is expressed as the mean proportional reduction of individual exposure behaviours, a steady-state flattened curve is achieved at an effective protective coverage of only 50% (Figure 1B and C). Even very modest further reductions beyond this threshold result in remarkably steep expected downslopes for epidemic contraction (Figure 1C). For example, incidence rates are expected to drop by a third every three weeks at an effective protective coverage of 60% (*R*_0_=0.64) and by almost two thirds (*R*_0_=0.36) at an effective protective coverage of 70% (Figure 1B and C).

Many non-specialists are familiar with the explosive dynamics of exponential growth, reflected in the steep upward trajectories expected for 5% shortfalls relative to *flatten the curve* containment targets (Figure 1A and C). However, the equally important implications of non-linearities in exponential decay curves are less widely appreciated. Analogous to eating a cake in proportion to its remaining size, it takes a long time to get down to the last crumbs if one eats a third, and then a third of the remaining two thirds, and so on. By comparison, consuming two thirds the first time, and then two thirds of the remaining one third the second time, reduces the cake much faster. In fact, the remaining fraction of the hypothetical cake will be four times smaller (1/9 versus 4/9) after only removing two slices and the difference in relative size grows rapidly as these two trajectories proceed. The same simple arithmetic rules apply to epidemic containment, so the expected trajectories for 60% and 70% effective protective coverage in figure 1C resemble those for our hypothetical cake. Correspondingly, these two modestly ambitious containment levels, which differ by only 10% in terms of effective protective coverage, need to be maintained for very different periods before the last cases are expected to occur. While lowering the incidence rate from 10,000 to only 1 case per week is expected to take 60 weeks at 60% effective protective coverage, the same near-elimination threshold would be reached after only 27 weeks at 70% effective protective coverage (Figure 1C). At 80% effective protective coverage, only 15 weeks are required to approach elimination (Figure 1C), and while the “10 weeks to crush the curve” hypothesis^7^ appears questionably optimistic, it might nevertheless be plausible if 85% protective coverage could be achieved.

Furthermore, the rapid growth of expected incidence rates for *flatten the curve* strategies that fall only 5% short of their targets (Figure 1A and C) underlines the fundamental dangers of this approach. It also highlights the fact that there is very little room for relaxing current restrictions in many countries where they have proven barely sufficient to contain the epidemic and begin slowly shrinking it.^11^ Considering how easily and rapidly epidemics may spiral out of control when restrictions are relaxed or viral reproduction surges for a variety of other reasons (Figure 1A and C), it is vital to remember that tipping points tip in both directions and are therefore dangerous places to linger. Deliberately planning to establish near-steady-state equilibria for epidemics with naturally volatile dynamics that are difficult to predict^9^ is risky at best. Additional risks of allowing SARS-CoV-2 transmission to continue include indefinite persistence among humans through unstable endemic transmission,^9^ establishment of zoonotic reservoirs, and rapid evolution of a large viral population into new forms that could be even more difficult to contain.

The ambitious containment and exclusion requirements of *crush the curve* strategies are obviously substantive undertakings. Success will require meticulous closure of remaining gaps in preventative intervention coverage, as well as comprehensive containment of case importation through travel and trade.^2^ However, many tractable opportunities remain to be exploited for closing the various intervention loopholes that allow residual transmission to persist through essential workers, goods and services of all kinds.^2^ And many encouraging precedents exist for certifying countries as free from infection with veterinary pathogens like swine fever^12^ or human pathogens like malaria.^13^ It is also encouraging that viral outbreaks of Ebola in 2014 and 2018, Severe Acute Respiratory Syndrome in 2003 and Middle East Respiratory Syndrome in 2012 all threated to become larger pandemics but were successfully contained and eliminated. However, the most convincing reason to be optimistic about SARS-CoV-2 is that several countries in Asia and the Pacific that have already crushed their epidemic curves are well on the way to elimination and exclusion endpoints.

More to the point, there appears to be no other safe and sensible option going forward that doesn’t necessitate extending most existing restrictions and their inevitable socioeconomic consequences for years, if not indefinitely.^8,9^ And as in any competitive sport, playing a defensive game against an unpredictable, fast- moving, adaptable and unrelenting opponent over a long drawn out game is asking for trouble.

## Data Availability

All models and data for this study are provided as a supplementary file.

## Notes

### Competing Interest Statement

The authors have declared no competing interest.

### Funding Statement

No funding support was received for this article

